# Exploratory Study on COPD Phenotypes and their Progression: Integrating SPECT and qCT Imaging Analysis

**DOI:** 10.1101/2024.04.10.24305577

**Authors:** Frank Li, Xuan Zhang, Alejandro P. Comellas, Eric A. Hoffman, Michael M. Graham, Ching-Long Lin

**Affiliations:** Roy J. Carver Department of Biomedical Engineering, University of Iowa, Iowa City, Iowa, USA; IIHR-Hydroscience & Engineering, University of Iowa, Iowa City, Iowa, USA; Department of Mechanical Engineering, University of Iowa, Iowa City, Iowa, USA; Department of Internal Medicine, University of Iowa, Iowa City, Iowa, USA; Department of Radiology, University of Iowa, Iowa City, Iowa, USA

**Keywords:** CT, SPECT, COPD, Ventilation, Small Airway Disease

## Abstract

**Background:** The objective of this study is to understand chronic obstructive pulmonary disease (COPD) phenotypes and their progressions by quantifying heterogeneities of lung ventilation from the single photon emission computed tomography (SPECT) images and establishing associations with the quantitative computed tomography (qCT) imaging-based clusters and variables.

**Methods:** Eight COPD patients completed a longitudinal study of three visits with intervals of about a year. CT scans of these subjects at residual volume, functional residual capacity, and total lung capacity were taken for all visits. The functional and structural qCT-based variables were derived, and the subjects were classified into the qCT-based clusters. In addition, the SPECT variables were derived to quantify the heterogeneity of lung ventilation. The correlations between the key qCT-based variables and SPECT-based variables were examined.

**Results:** The SPECT-based coefficient of variation (CV_Total_), a measure of ventilation heterogeneity, showed strong correlations (|r| ≥ 0.7) with the qCT-based functional small airway disease percentage (fSAD%_Total_) and emphysematous tissue percentage (Emph%_Total_) in the total lung on cross-sectional data. As for the two-year changes, the SPECT-based maximum tracer concentration (TC_max_), a measure of hot spots, exhibited strong negative correlations with fSAD%_Total_, Emph%_Total_, average airway diameter in the left upper lobe, and airflow distribution in the middle and lower lobes.

**Conclusion:** Small airway disease is highly associated with the heterogeneity of ventilation in COPD lungs. TC_max_ is a more sensitive functional biomarker for COPD progression than CV_Total_. Besides fSAD%_Total_ and Emph%_Total_, segmental airways narrowing and imbalanced ventilation between upper and lower lobes may contribute to the development of hot spots over time.

## INTRODUCTION

Chronic obstructive pulmonary disease (COPD) is a prevalent respiratory disease that imposes a significant burden on healthcare systems worldwide (1). The Global Initiative for Chronic Obstructive Lung Disease (GOLD) report (2) has been continuously updated to better characterize COPD heterogeneities and improve treatment outcomes (3,4). In addition, imaging modalities, such as computed tomography (CT) and single-photon emission computed tomography (SPECT), have been utilized to visualize and quantify regional functional and structural changes associated with various pathological processes in the lung diseases such as emphysema (5).

Innovative biomarkers based on quantitative CT (qCT) imaging have been developed to investigate the underlying causes of COPD. These biomarkers assess a wide range of risk factors and associated defects, such as airway-branch variation (6), dysanapsis (7), and pulmonary vascular dysfunction (8,9). The introduction of the qCT-based parametric response map (PRM) (10) has also significantly advanced research on functional small airway disease (fSAD) and emphysema (Emph). Both fSAD and Emph are critical phenotypes in COPD, with the former considered a precursor to the latter (11). It is suggested that targeting small airways with appropriate treatments may potentially control the progression of both airway and parenchymal diseases in COPD (12). Novel multiscale qCT biomarkers have been developed to capture a wide range of COPD phenotypes at different disease stages. These imaging-based biomarkers allow for identification of clusters (also known as subtypes, subgroups, or subpopulations) of COPD patients that show strong associations with clinical characteristics. For instance, Haghighi et al. applied unsupervised clustering techniques to cross-sectional qCT biomarkers and identified four clinically-relevant imaging-based clusters among former and current smokers, respectively, to eliminate the effect of inflammation on the emphysema index (13,14). These clusters were sorted in order of severity, with cluster 1 (C1) being the mildest and cluster 4 (C4) being the most severe in terms of airflow obstruction. Classification of COPD patients into imaging-based clusters can aid in comprehending the heterogeneous nature of the disease more quantitatively and locally. In addition, Zou et al. utilized the multiscale qCT phenotyping and clustering mentioned above to analyze longitudinal data from two visits to identify COPD progression clusters (15). Cross-sectional clustering is based on static severity stages or phenotypes, while longitudinal clustering aims to capture disease activity mechanisms or endotypes.

Assessment of global and regional lung ventilation can be achieved through SPECT ventilation imaging, which measures the concentration of a radioactive tracer serving as a biomarker for ventilation if the tracer aerosol size is small enough to reach alveoli. De Backer et al. demonstrated a strong correlation between CT-based lobar air volume change and lobar SPECT tracer aerosol concentration with aerosol diameter less than 2µm (16). They further demonstrated that the hot spots observed on SPECT images corresponded to airway constrictions on CT images as well as the hot spots determined by computational fluid dynamics simulation. Studies have also demonstrated a correlation between heterogeneities observed on SPECT images and impaired lung functions. Xu et al. found that the coefficient of variation (CV) observed in SPECT images, which is a measure of ventilation heterogeneity, not only differentiated patients with emphysema from non-emphysematous smokers and non-smokers, but also exhibited a correlation with pulmonary functions. (17). Thus, the use of combined SPECT and CT imaging enables the evaluation of the functional and structural relationships within different regions of the lungs (5). Recently approved by the FDA, hyperpolarized xenon-129 magnetic resonance imaging (XeMRI) can serve as an alternative approach to evaluate lung ventilation distribution without exposing patients to ionizing radiation (18–21).

In this study, we aimed to evaluate COPD progression by quantifying structural and functional changes in CT and SPECT images of COPD subjects acquired at three visits, with approximately one year between each visit. Our objective was to establish the links between qCT variables and SPECT biomarkers within the previously established COPD qCT-based clusters (13,14). Given the sample size in this study, the associations observed with the imaging clusters may assist with our understanding of COPD progression. To the best of our knowledge, this study was the first attempt to conduct a longitudinal CT/SPECT analysis, shedding light on COPD progression.

## METHOD

Fourteen COPD patients were recruited to participate in a longitudinal study of three visits at 0, 12.84 ± 1.68, and 26.20 ± 3.06 months, denoted by V0, V1, and V2, respectively. Five subjects dropped out of the study due to other health issues or personal reasons, and one subject was excluded due to an abnormal airway structure having an accessary bronchus connected to the right main bronchus. Thus, eight subjects who completed the three visits were analyzed in this study. Pulmonary function tests (PFT), CT scans, and SPECT scans were acquired at all visits. This study was approved by the institutional review board of the University of Iowa and the informed consent was obtained from all the patients before the study.

### CT Images and qCT Variables

Three static three-dimensional (3D) CT scans were acquired using the Siemens SOMATOM Force Scanner at the lung volumes of total lung capacity (TLC), functional residual capacity (FRC) and residual volume (RV), using the lung volume control system proposed in the previous study (22). The TLC scan protocol used dose modulation with 36 reference mAs, 120 kV, pitch of 1.0, and rotation time of 0.25 s. The FRC and RV scans used dose modulation with 15 reference mAs, 120 kV, pitch of 1.0, and rotation time of 0.25 s The CT images have a spatial resolution of 0.5 mm in the z axis, and 0.5-0.7 mm in the x and y axes. The subjects were positioned on the CT table in the supine position and were instructed to breathe normally through the mouthpiece for a few breathing cycles. While the CT scans were performed, they were instructed to take in a deepest breath and hold it at the desired lung volume of TLC, FRC, or RV. The subject breathed normally for a few breathing cycles before the next CT scan was performed.

The CT images were processed by VIDA Vision (VIDA Diagnostics Inc., Coralville, Iowa) to segment the lung, lobes, and central airways. A mass preserving image registration technique was performed to match FRC (or RV) with TLC images (23). TLC and RV images were registered to derive functional variables, while TLC and FRC images were registered to match SPECT images. A total of 69 structural and functional multiscale qCT variables presented in previous studies were derived (13–15) as shown in **Figure 1**. The structural and functional variables describe the regional alterations of lung structures and capture the regional alterations of lung functions, respectively. The qCT variables most relevant to this study include normalized airway wall thickness (WT*), normalized airway hydraulic diameter (D_h_*), fractional air volume change (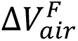), determinant of Jacobian matrix (J), anisotropic deformation index (ADI), fraction-based small airways disease (fSAD%), fraction-based emphysema (Emph%), and tissue fraction at TLC (β_tissue_). Please refer to **Supplementary Material** for detailed explanation of these variables and a list of abbreviations of qCT variables. Subsequently, the subjects were classified into pre-established imaging-based clusters (13,14) using these qCT variables, aiming to enhance our understanding of COPD progression within a broader population context. The assignment of cluster membership was determined based on the distances to the geometric centers of the clusters within the qCT feature space.

**Figure 1.**
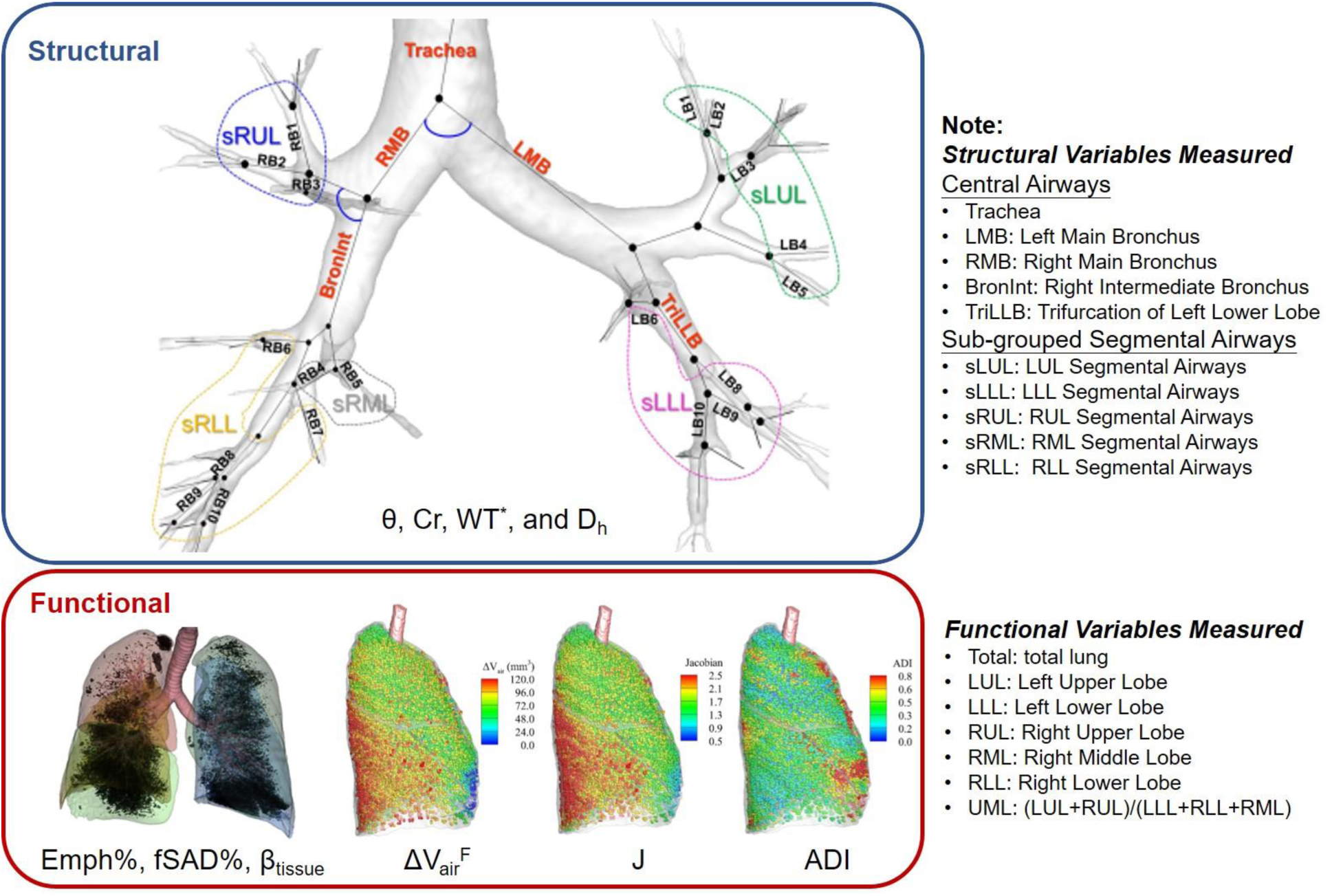
The multiscale structural and functional qCT variables.

The region, where the qCT variable was measured, was specified as the subscript of the variable, namely {Variable}_{Region}_. The structural variables were measured from the lung shape, central airways, and segmental airways. The lung shape was measured at TLC as the ratio of apical-basal distance and ventral-dorsal distance. The central airways include trachea, left main bronchus (LMB), right main bronchus (RMB), right intermediate bronchus (BronInt), and trifurcation of left lower lobe (TriLLB). The segmental airways were grouped by lobes, such as sub-grouped segmental airways of left upper lobe (sLUL), sub-grouped segmental airways of left lower lobe (sLLL), sub-grouped segmental airways of right upper lobe (sRUL), sub-grouped segmental airways of right middle lobe (sRML), and sub-grouped segmental airways of right lower lobe (sRLL). The functional variables were measured at lobar level, including left upper lobe (LUL), left lower lobe (LLL), right upper lobe (RUL), right middle lobe (RML), and right lower lobe (RLL), as well as total lung level (Total).

### SPECT Images and Variables

The subject was positioned in the supine position on the SPECT scanner to duplicate the same position used in the above CT scans. The patient inhaled through a mouthpiece, with the nose occluded, ^99m^Tc sulfur colloid as a radioactive tracer aerosol that was generated by a specialized nebulizer. Then a 3D SPECT scan of the whole lung was performed. The particle size of the sulfur colloid is below 1 µm (24), enabling distribution of the tracer aerosol deep into the lung (25). Since ^99m^Tc sulfur colloid is non-absorbable, it is not cleared from the lung rapidly. The 3D SPECT scan took approximately 20 minutes to complete and was acquired during continuous tidal breathing. The SPECT images have a spatial resolution of 3.895 mm in the x, y, and z axes. It was estimated that the subject retained a dose of 0.17 mSv, which was lower than the standard of the U.S. Nuclear Regulatory Commission (50 mSv/year). It is noted that the original SPECT imaging protocol was to use ^99m^Tc-HMPAO labeled neutrophils to evaluate the distribution of inflammation, and was adopted for the first two subjects at V0. However, the resulting SPECT images did not provide the expected results. We then modified the protocol. Thus, the first two subjects did not have SPECT ventilation scans at V0.

SPECT image is a dynamic scan which is a time-averaged image acquired during tidal breathing. On the other hand, the CT images are static scans acquired at TLC, FRC, and RV. It is preferable to match the SPECT images with the FRC CT images, which were taken at a lung volume closer to tidal breathing, than with the RV or TLC CT images. The lung deformation from FRC to the peak of tidal volume was assumed relatively small and quasi-linear. Thus, the SPECT images were first matched with the FRC CT images by affine transforms, using mutual information as the cost function. Displacement fields of transformation from FRC to TLC images were obtained by image registration. Subsequently, the transformed SPECT images were deformed by applying the displacement fields to match the TLC CT images, which provide detailed anatomical information of the lungs.

After matching the SPECT images with TLC CT images, the tracer concentrations (TC) within the lung and each lobe were quantified at the domain of TLC CT images. The ratio of TC in a lobe over the total lung (TC%), TC coefficient of variation of the total lung (CV_Total_), and standardized maximum TC (TC_Max_) in the total lung were used to describe the distribution, heterogeneity, and hot spot of TC. Specifically, TC% was used to measure the lobar ventilation distribution. CV_Total_ was a standardized measure of the heterogeneity of TC distribution in the total lung, calculated by normalizing the standard deviation with the average of TC (17). TC_Max_ was used to quantify the local hot spots magnitude as a measure of airway constriction. The normalized CV_Total_ and TC_Max_ aimed to factor out the effect of different amount of TC inhaled. Moreover, the correlation between TC% and 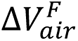 was used to assess the efficacy of the image registration of SPECT and CT images. The Pearson’s correlation coefficients between CV_Total_ and PFT values were calculated and validated against previous studies (17,26).

### Analysis of SPECT and qCT Variables

The associations between SPECT and qCT variables were evaluated to assist the interpretation of the SPECT variables. Exploratory factor analysis (EFA) was applied to reduce the large number of qCT variables derived from the 799 current and former smokers (13,14) to a lower number of factors that preserved the variability of qCT-captured features. The number of factors was determined by the parallel analysis (27). The factors were extracted by principal component analysis (PCA) with Varimax rotation (28). Varimax rotation is an orthogonal rotation, so the factors are independent.

We then selected key qCT variables contributing significantly to each factor as the surrogates of the factors for interpretability of disease phenotypes. The qCT variables were regarded as key variables if their loading values on their corresponding factors were greater than 0.7. We then examined the correlations between key qCT variables and SPECT variables, based on the cross-sectional data at respective V0, V1, and V2 as well as the longitudinal data with changes between any two visits of V1-V0, V2-V1, and V2-V0. Since this study was exploratory and the sample size was small, we only focused on the large effect size, specifically the Pearson’s correlations with coefficient greater than 0.7 (29). A large effect size indicates that even with a small sample size, the observed effect is likely to be substantial and meaningful.

## RESULT

Five former smokers (Subject 1-5) and three current smokers (Subject 6-8) were analyzed (age: 63.1 ± 11.5 years; range: 51-84 years). The cluster characteristics of former smokers and current smokers were summarized in **Table 1**. The cluster membership, clinical data, and PFTs of these subjects were presented in **Table 2** (see **Table S1** for more data). The subjects had an average of Tiffeneau ratio (FEV_1_/FVC%) of 52.9 ± 16.6 %, and an average of predicted forced expiratory volume ratio (FEV_1_% predicted) of 67.9 ± 16.3% at V0. The mean changes between V1 and V0 in terms of FEV_1_/FVC% and FEV_1_% predicted were 0.0 ± 1.9% and −2.5 ± 5.4%, respectively. Moreover, the mean changes between V2 and V1 in terms of FEV_1_/FVC% and FEV_1_% predicted were 0.8 ± 1.9% and 3.5 ± 4.6%, respectively.

**Table 1.** Characteristics of clusters of the current and former smokers, defined by Haghighi et al. (13,14).

|  | Current Smokers | Former Smokers |
| --- | --- | --- |
| **C1** | Relatively resistant smokers with preserved pulmonary function | Asymptomatic resistant smokers with preserved pulmonary function |
| **C2** | Airway-wall-thickening; fSAD-dominant subjects with obesity and activity limitation | Obese female individuals with preserved lung function and marginal emphysema |
| **C3** | Airway-wall-thinning; fSAD-dominant subjects | Older male individuals with increasing fSAD and emphysema |
| **C4** | Severe emphysema-fSAD-mixed subjects with severe airway luminal narrowing and wall thinning | Severe emphysema and fSAD individuals with severe structural alterations |

**Table 2.** Clinical data, PFT at V0, and changes in terms of PFT between visits. Subject 1-5, former smokers; Subject 6-8, current smokers.

| Subject | Gender | Age (yrs.) | BMI | GOLD/Cluster | V0 |  | V1 – V0 |  | V2 – V0 |  |
| --- | --- | --- | --- | --- | --- | --- | --- | --- | --- | --- |
|  |  |  |  |  | FEV<sub>1</sub>% predicted | FEV<sub>1</sub>/FVC % | FEV<sub>1</sub>% predicted | FEV<sub>1</sub>/FVC % | FEV<sub>1</sub>% predicted | FEV<sub>1</sub>/FVC % |
| 1 | M | 71-80 | 32 | 2/3 | 73 | 53 | 1 | 3 | 2 | -3 |
| 2 | M | 81-90 | 23 | 2/4 | 50 | 28 | -11 | -2 | 12 | 3 |
| 3 | M | 51-60 | 26 | 2/1 | 63 | 51 | -2 | 0 | 1 | 0 |
| 4 | M | 51-60 | 21 | 2/3 | 73 | 56 | -5 | 2 | 10 | 2 |
| 5 | M | 51-60 | 32 | 3/3 | 36 | 28 | -5 | -3 | 2 | 3 |
| 6 | M | 51-60 | 29 | 1/1 | 83 | 59 | -5 | -1 | 3 | 1 |
| 7 | F | 51-60 | 27 | 0/3 | 77 | 71 | -2 | 0 | -2 | 0 |
| 8 | F | 61-70 | 36 | 0/1 | 88 | 77 | 9 | 1 | 0 | 0 |
| **Median (Range)** |  |  | 28(21-36) |  | 73 (36 - 88) | 54.5 (28 - 77) | -3.5 (-11 - 9) | 0 (-3 - 3) | 2 (-2 - 12) | 0.5 (-3 - 3) |

### SPECT Quantification

TC% and 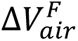 of each lobe at each visit were listed in **Table S2** of the supplementary material. The correlation coefficient between TC% and 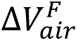 was 0.73, indicating a strong correlation between the two variables (**Figure 2 (a)**). In addition, the correlations between CV_Total_ and PFT values showed a strong negative correlation with FEV_1_% predicted at −0.74 and an even stronger negative correlation with FEV_1_/FVC% at −0.80 (**Figure 2 (b)**).

**Figure 2.**
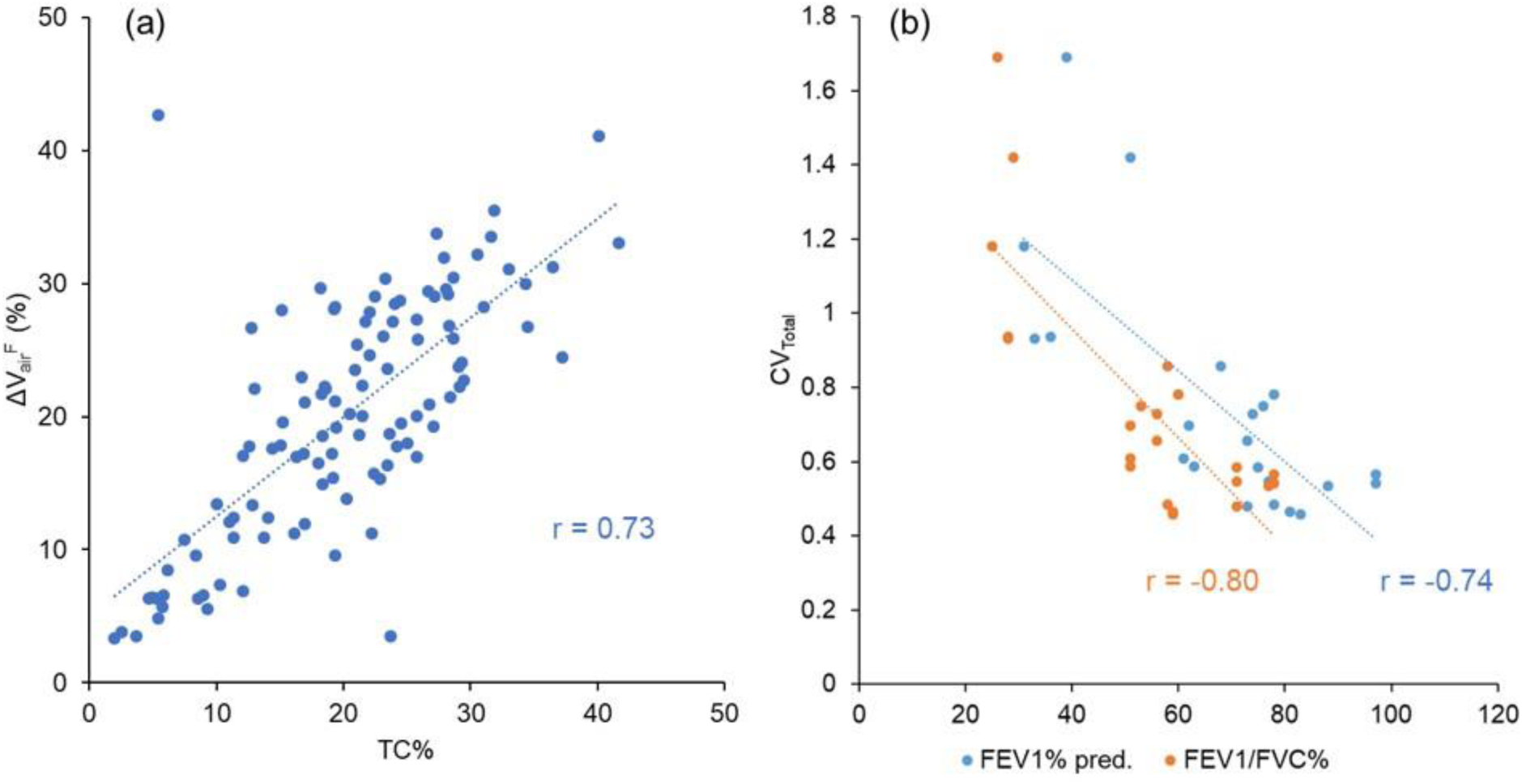
(a) A scatter plot showing the strong correlation (r=0.73) between TC% and ΔV_air_^F^ of the lobes. (b) A scatter plot showing the strong correlations between CV_Total_ and PFT results (FEV_1_% predicted: r=-0.74, FEV_1_/FVC%: r=-0.80). The data points were from both three visits.

The SPECT images for all the subjects were displayed in **Figure 3**. CV_Total_ and TC_Max_ at each visit together with their changes between visits were shown in **Table 3**. At all the visits, Subject 2 and 5 were characterized by greater CV_Total_ and extremely high TC_Max_, while Subject 1, 6, 7, and 8 exhibited more homogeneous TC distributions. Subject 3 and 4 had large TC_Max_ at V1 and V2, respectively. Hot spots revealed by concentrated red areas were notable in the SPECT images of Subject 2 and 5 for all the visits. A distinguished hot spot was also observed in the SPECT image of Subject 3 at V2. The changes of CV_Total_ and TC_Max_ between visits for all the subjects were small, being around 10%.

**Figure 3.**
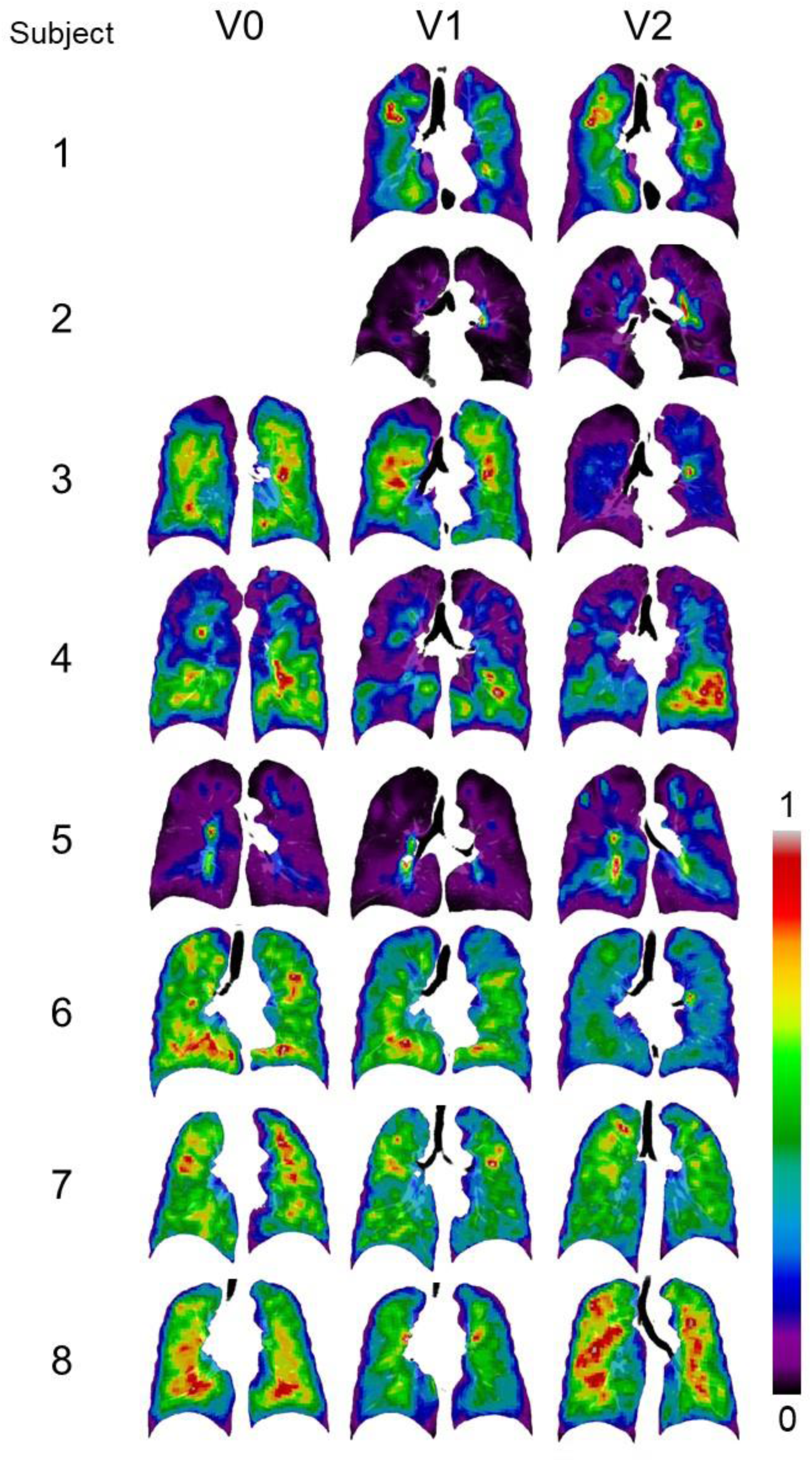
Normalized SPECT images transformed to TLC domain at the three visits. Subject 1 and 2 did not have ventilation scans at V0.

**Table 3.** CV_Total_ and TC_Max_ at each visit, and their changes between visits.

| Subject | V0 | V1 | V2 | V1-V0 | V2-V1 | V2-V0 |
| --- | --- | --- | --- | --- | --- | --- |
| **CV<sub>Total</sub>** |  |  |  |  |  |  |
| 1 |  | 0.73 | 0.75 |  | 0.02 |  |
| 2 |  | 1.69 | 1.42 |  | -0.27 |  |
| 3 | 0.59 | 0.61 | 0.7 | 0.02 | 0.09 | 0.11 |
| 4 | 0.66 | 0.86 | 0.78 | 0.2 | -0.08 | 0.13 |
| 5 | 0.94 | 1.18 | 0.93 | 0.24 | -0.25 | 0 |
| 6 | 0.46 | 0.48 | 0.47 | 0.03 | -0.02 | 0.01 |
| 7 | 0.55 | 0.58 | 0.48 | 0.04 | -0.1 | -0.07 |
| 8 | 0.53 | 0.57 | 0.54 | 0.03 | -0.02 | 0.01 |
| **Median (Range)** | 0.57 (0.46 - 0.94) | 0.67 (0.48 - 1.69) | 0.725 (0.47 - 1.42) | 0.035 (0.02 - 0.24) | -0.05 (-0.27 - 0.09) | 0.01 (-0.07 - 0.13) |
| **TC<sub>Max</sub>** |  |  |  |  |  |  |
| 1 |  | 7.27 | 5.36 |  | -1.91 |  |
| 2 |  | 37.69 | 20.83 |  | -16.86 |  |
| 3 | 7.32 | 5.36 | 16.33 | -1.97 | 10.97 | 9.01 |
| 4 | 5.03 | 14.22 | 7.82 | 9.19 | -6.4 | 2.79 |
| 5 | 24.1 | 18.66 | 19.59 | -5.44 | 0.93 | -4.51 |
| 6 | 3.51 | 4.19 | 9.23 | 0.68 | 5.05 | 5.72 |
| 7 | 4.14 | 5 | 4.2 | 0.86 | -0.81 | 0.06 |
| 8 | 3.82 | 4.21 | 3.55 | 0.39 | -0.65 | -0.26 |
| **Median (Range)** | 4.585 (3.51 - 24.1) | 6.315 (4.19 - 37.69) | 8.525 (3.55 - 20.83) | 0.535 (-5.44 - 9.19) | -0.73 (-16.86 - 10.97) | 1.425 (-4.51 - 9.01) |

### Subject Classification based on qCT Variables

At V0, Subject 1-5 were classified into respective cluster 3, 4, 1, 3, and 3 (C3, C4, C1, C3, and C3), of former-smoker clusters (14), respectively. The memberships of these subjects did not change from V0 to V1. However, from V1 to V2, Subject 1 changed from C3 to C1, Subject 5 moved from C3 to C4, and Subject 2-4 remained the same clusters. On the other hand, Subject 6, 7, and 8 were classified into C1, C3, and C1 of current-smoker clusters (13), respectively, at both V0 and V1. However, from V1 to V2, Subject 7 changed from C3 to C1 and Subject 6 and 8 remained the same clusters. In summary, among these 24 imaging-based classifications (for 8 subjects at 3 visits), only three changed at V2, indicating the stability of cluster membership over two years. These classifications were placed on the space formed by the first and the second principal components (PC0 and PC1) to visualize the progressive loci of these subjects in the cluster landscape depicted by the cohorts of former and current smokers studied previously (13,14) (**Figure 4**). The imaging metrics that contributed the most to PC0 were fSAD%_Total_ and Emph%_Total_, while the imaging metrics that contributed the most to PC1 were β_tissue_,_Total_, J_Total_, and ADI_Total_ (**Figure S1**). Subject 1 (former smoker) and Subject 7 (current smoker) displayed the similar loci, moving downward along PC1 and changing from C3 to C1 at V2, while Subject 5 moving rightward along PC0 and changing from C3 to C4 at V2. Subject 2, 3, 4, 6, and 8 remained within their respective cluster domains during the two-year duration.

**Figure 4.**
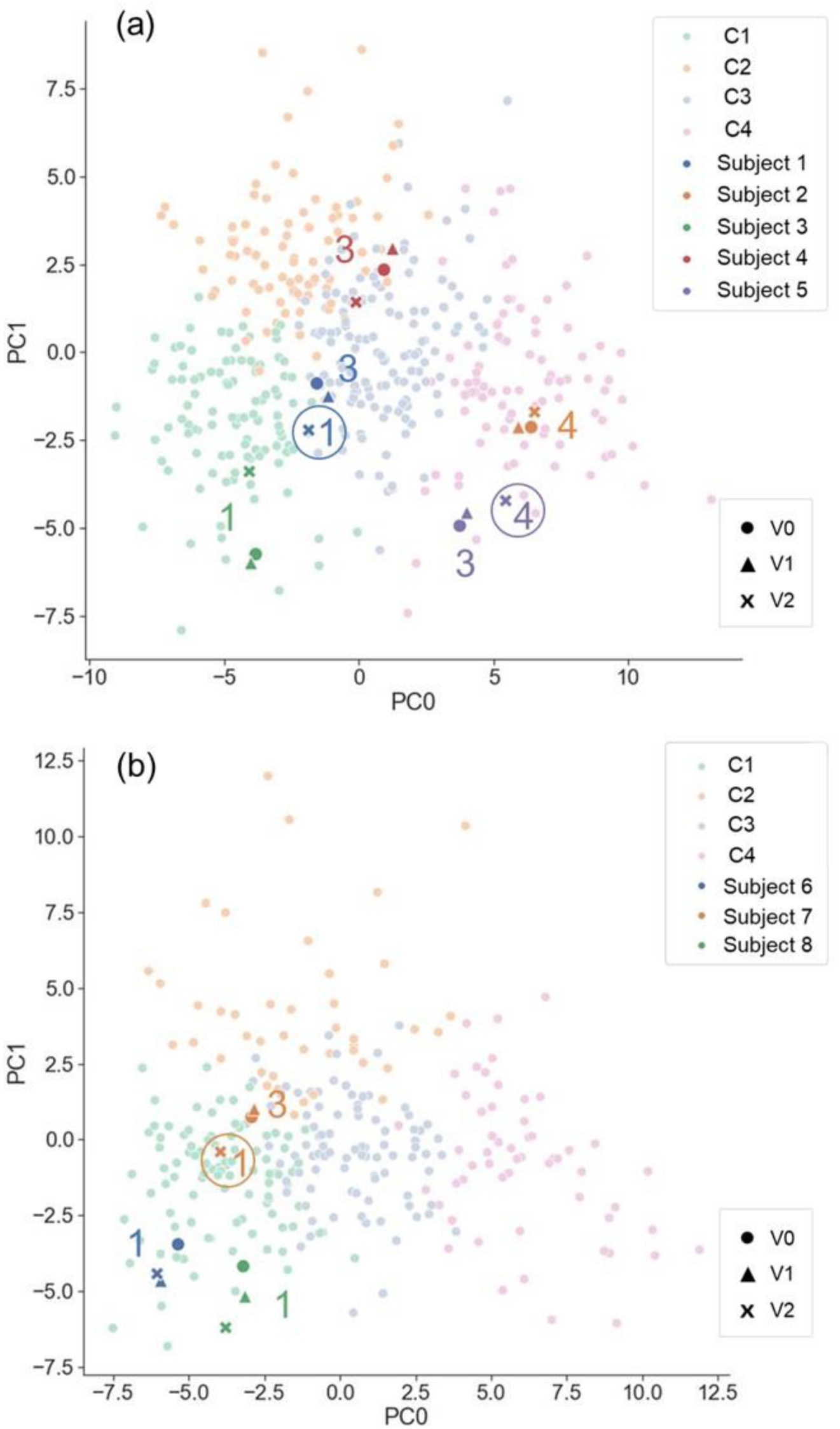
(a) Former smokers in this study placed on the two-dimensional space formed by the first and the second principal components derived from previous study (14). (b) Current smokers in this study placed on the two-dimensional space formed by the first and the second principal components derived from previous study (13). C1, C2, C3, and C4 in the figure legend denote all of the subjects analyzed in previous studies (13,14). Those enclosed by a circle denote the change of cluster membership.

### Factors Extracted from Former and Current Smokers

Seven factors were extracted from the qCT variables of the former and current smokers (13,14). The factors were able to explain 78.4% of the variance of the original features. The important qCT variables which had moderate-to-high and high contribution (factor loading > 0.6) to the factors were listed in **Table 4**. ADI_Total_ and J_Total_ were selected to represent Factor 0 (F0). β_tissue_,_Total_ and fSAD%_Total_ were selected to represent Factor 1 (F1). D_h_*_sLUL_ was selected to represent Factor 2 (F2). Emph%_Total_ was selected to represent Factor 3 (F3). 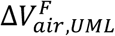 was selected to represent Factor 4 (F4). WT*_sLLL_ was selected to represent Factor 5 (F5). D_h_*_LMB_ was selected to represent Factor 6 (F6).

**Table 4.** The important qCT variables which have large contribution to the factors. The variables shown in this table had the magnitude of factor loadings greater than 0.6. A “(-)” indicates a negative factor loading.

| **F0** | **F1** | **F2** | **F3** | **F4** | **F5** | **F6** |
| --- | --- | --- | --- | --- | --- | --- |
| ADI<sub>Total</sub> | $\beta_{\text{tissue, Total}}$ | D<sub>h</sub><sup>*</sup><sub>sLUL</sub> | Emph%<sub>Total</sub> | $\Delta V_{\text{air,UML}}^F$ | WT<sup>*</sup><sub>sLLL</sub> | D<sub>h</sub><sup>*</sup><sub>LMB</sub> |
| J<sub>Total</sub> | $\beta_{\text{tissue, LLL}}$ | D<sub>h</sub><sup>*</sup><sub>sRLL</sub> | Emph%<sub>LUL</sub> | $\Delta V_{\text{air,LUL}}^F$ | WT<sup>*</sup><sub>sRUL</sub> | WT<sup>*</sup><sub>LMB</sub> |
| J<sub>RLL</sub> | $\beta_{\text{tissue, RLL}}$ | D<sub>h</sub><sup>*</sup><sub>sLLL</sub> | Emph%<sub>RUL</sub> | $\Delta V_{\text{air,RUL}}^F$ | WT<sup>*</sup><sub>sRLL</sub> | D<sub>h</sub><sup>*</sup><sub>RMB</sub> |
| J<sub>RUL</sub> | $\beta_{\text{tissue, LUL}}$ | D<sub>h</sub><sup>*</sup><sub>TriLLB</sub> | Emph%<sub>RLL</sub> | $\Delta V_{\text{air,RLL}}^F$ (-) | WT<sup>*</sup><sub>TriLLB</sub> | |
| J<sub>LUL</sub> | $\beta_{\text{tissue, RML}}$ | D<sub>h</sub><sup>*</sup><sub>sRUL</sub> | Emph%<sub>LLL</sub> | $\Delta V_{\text{air,LLL}}^F$ (-) | WT<sup>*</sup><sub>sLUL</sub> | |
| J<sub>LLL</sub> | $\beta_{\text{tissue, RUL}}$ | D<sub>h</sub><sup>*</sup><sub>sRML</sub> | Emph%<sub>RML</sub> | | WT<sup>*</sup><sub>sRML</sub> | |
| ADI<sub>LUL</sub> | fSAD%<sub>RML</sub> (-) |  |  |  |  |  |
| J<sub>RML</sub> | fSAD%<sub>Total</sub> (-) |  |  |  |  |  |
| ADI<sub>RUL</sub> | fSAD%<sub>LUL</sub> (-) |  |  |  |  |  |
| ADI<sub>RLL</sub> | fSAD%<sub>RUL</sub> (-) |  |  |  |  |  |
| ADI<sub>LLL</sub> | fSAD%<sub>RLL</sub> (-) |  |  |  |  |  |
| ADI<sub>RML</sub> | fSAD%<sub>LLL</sub> (-) |  |  |  |  |  |

### Association between SPECT and qCT Variables

Association tests were conducted using both cross-sectional and longitudinal data. The cross-sectional tests captured inter-subject variation, while the longitudinal tests captured intra-subject progression. On cross-sectional data, both SPECT variables of CV_Total_ and TC_Max_ had the strongest correlations with fSAD%_Total_ and Emph%_Total_ among all qCT variables. The correlations of CV_Total_ with fSAD%_Total_ and Emph%_Total_ were 0.90 and 0.71, respectively. Similarly, the correlations of TC_Max_ with fSAD%_Total_ and Emph%_Total_ were 0.86 and 0.77, respectively. The strength of correlations between the one-year changes in SPECT variables and those in qCT variables was found to be weak. As for the two-year changes, TC_Max_ exhibited strong correlations with several qCT variables, including fSAD%_Total_ (r=-0.70), D_h,sLUL_* (r=-0.74), Emph%_Total_ (r=- 0.75), and 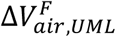 (r=0.75) (**Figure 5**). This finding suggests that TC_max_ is a more sensitive functional biomarker for COPD progression than CV_Total_.

**Figure 5.**
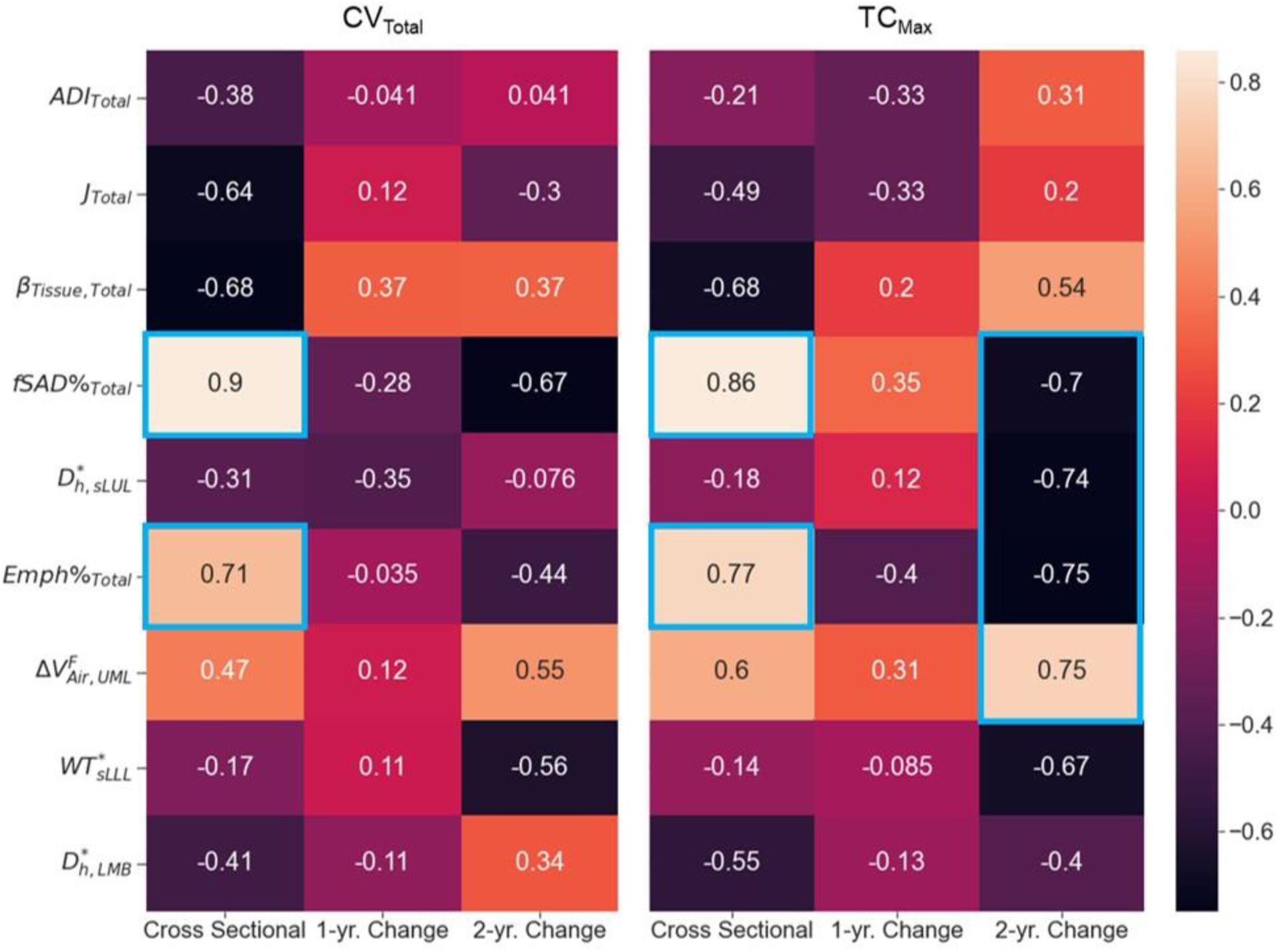
The correlations between the SPECT variables and the qCT variables for cross-sectional and longitudinal data. Magnitudes of correlations greater than 0.7 were bounded by blue boxes.

On longitudinal data, the selected qCT variables at each visit and their changes between visits were shown in **Table 5**. For all the visits, Subject 2 and 5 exhibited higher fSAD%_Total_ and Emph%_Total_ compared to the other subjects. Moreover, Subject 1, 3, and 4 had moderate fSAD%_Total_ and Emph%_Total_, while Subject 6-8 had low fSAD%_Total_ and Emph%_Total_. However, the changes of the selected key qCT variables between visits for all subjects were small. The fSAD maps at V1, which marked the fSAD-pixels on CT coronal slices of the subjects, were compared with the SPECT images in **Figure 6**. Visual inspection supported a correlation between qCT-based fSAD maps and SPECT-based ventilation patterns by groups of Subject (1,3,4), (2,5), and (6,7,8) with moderate, high, and low fSAD%_Total_, respectively.

**Figure 6.**
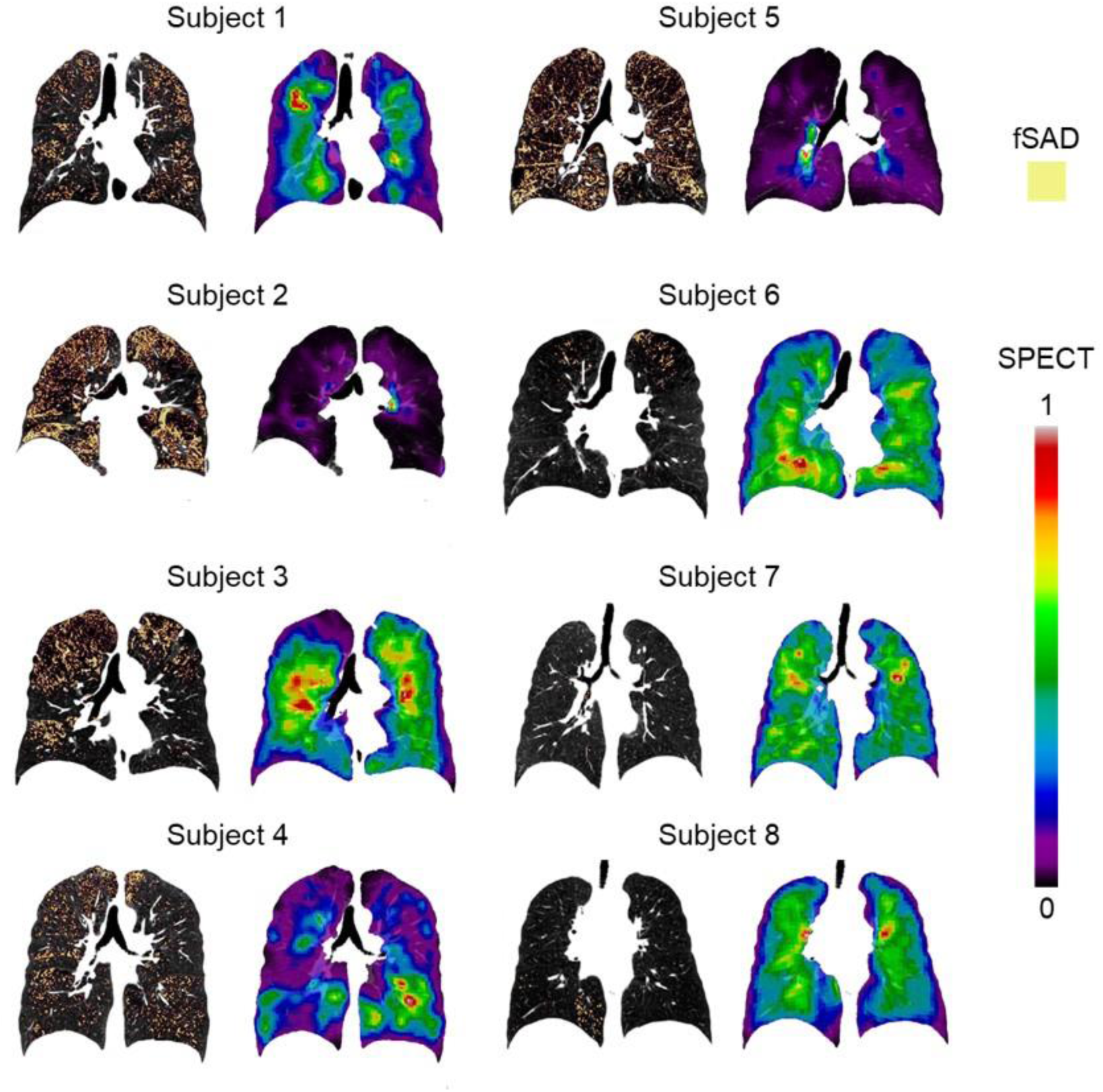
The fSAD-voxel maps for each subject (left column) along with their SPECT images (right column). These images were plotted in coronal planes of the CT images

**Table 5.**
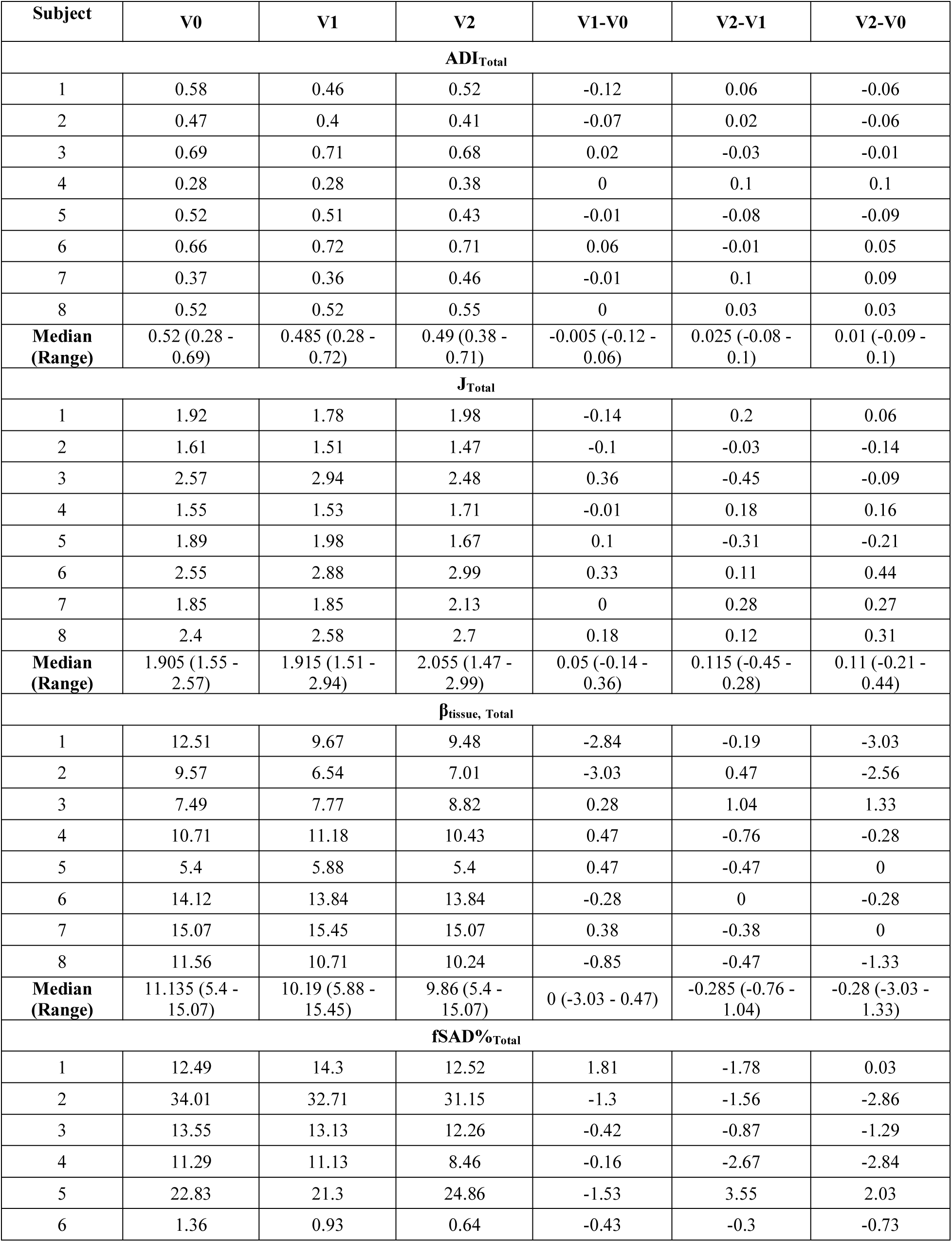
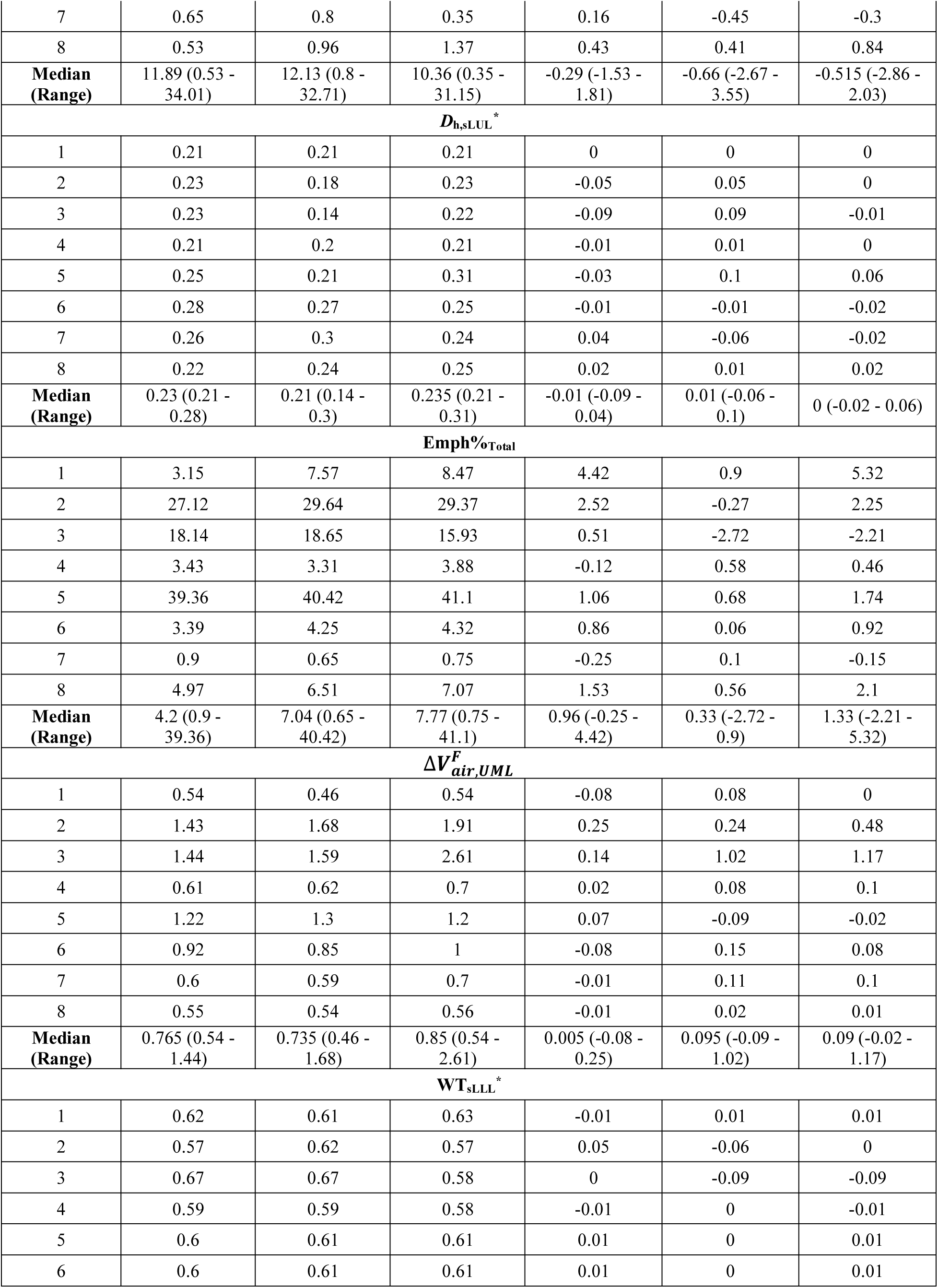
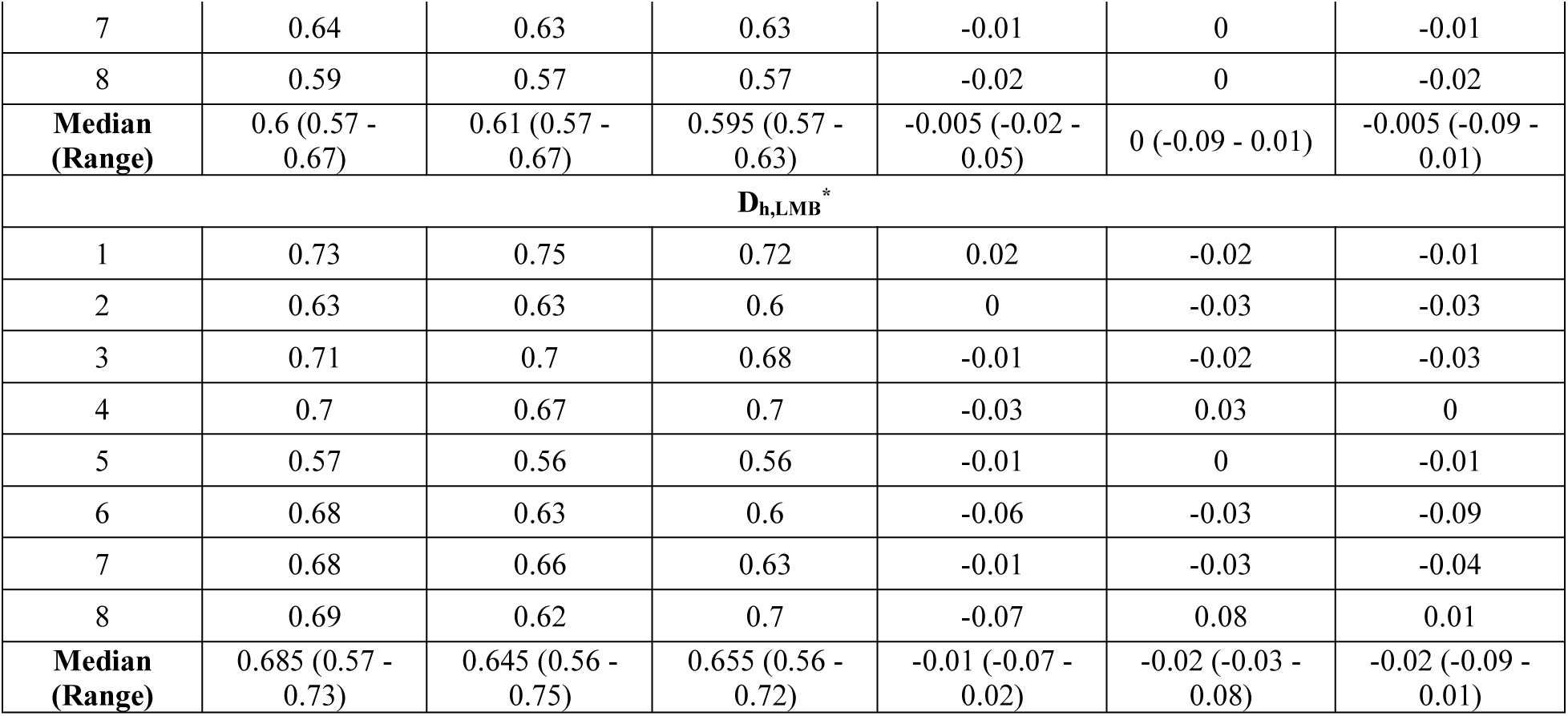
The selected qCT variables at each visit, and their changes between visits.

## DISCUSSION

In this work, five former smokers (Subject 1-5) and three current smokers (Subject 6-8) with COPD were studied using combined CT and SPECT imaging biomarkers. At V0, Subject 2 and 5 had moderate-to-severe (GOLD 2/3) and severe (GOLD 3) airflow obstruction, respectively. Subject 1, 3, 4, and 6 had either mild (GOLD 1) or moderate (GOLD 2) airflow obstruction. Subject 7 and 8 were regarded as subjects at risk of COPD (GOLD 0) according to their PFT results. The lung functions of all the subjects were well maintained over two years (**Table 2**).

The robust correlation observed between TC% and 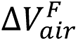 of the lobes (**Figure 2**: r=0.73) suggests that qCT-based 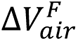 can be utilized as an alternative measure for lobar ventilation. The observed correlation also serves as evidence supporting the effectiveness of the registration process utilized to align SPECT and CT images. Furthermore, the notable negative correlations were found between TC heterogeneity (CV_Total_) and PFT results (**Figure 2**: r=-0.74 with FEV_1_% predicted; r=- 0.80 with FEV_1_/FVC%), agreeing with previous studies (r=-0.84 to -0.89 with FEV_1_% predicted) (17,18).

### Subject Classification based on qCT Variables

Subject 2 and 5 were patients with severe COPD. They had the most heterogeneous TC distribution with higher CV_Total_ and TC_Max_ compared to other subjects (**Figure 3**). The former-smoker clusters of C3 and C4 were characterized by elevated small airway disease and emphysema (**Table 1**). Subject 2 was classified into C4 for all the visits. Subject 5 was classified into C3 at V0, but progressed to C4 at V2, remaining near the boundary of C3 and C4 in the 2D latent space for all the visits (**Figure 4 (a)**). Thus, as expected, Subject 2 and 5 had greater fSAD%_Total_ and Emph%_Total_ than others.

Subject 1, 3, and 4 were COPD patients at moderate stages. Subject 1 and 4 were classified as C3 at V0 and V1. Although Subject 1 changed to C1 at V2, this subject remained near the boundary of C1 and C3 (**Figure 4 (a)**). Therefore, compared to C1 subjects, greater fSAD%_Total_ was observed, yet elevated Emph%_Total_ was not found for Subject 1 and 4 at all visits. This may explain why they were close to the boundaries of C1 and C2 and shared the characteristics of the two clusters, having marginal emphysema with relatively preserved lung function (14). It is noteworthy that the lung functions may not be sensitive enough to detect the elevated fSAD% (30). Due to the greater J_Total_ and less β_tissue, Total_ found in Subject 1 at V2, the cluster membership shifted from C3 to C1. Subject 1 and 4 showed moderate dispersion of the TC distribution, exhibiting more heterogeneous TC distribution than Subject 6-8 (**Figure 3**). Subject 3 was classified as C1 for all the visits due to large lung deformation from RV to TLC lung (as measured by J_Total_) regardless of the fact that Subject 3 had increased fSAD%_Total_ and Emph%_Total_. The large deformation might be attributable to the hyperinflated lung caused by emphysematous regions with less elastic air sacs (31). Since this subject was close to the boundary of C1 and C3 (**Figure 4 (a)**) and had elevated fSAD%_Total_ and Emph%_Total_, Subject 3 may be regarded as a C3 subject. The TC distributions for V0 and V1 were similar to those of Subject 1 and 4 at all visits. Furthermore, a clear hot spot was observed on the SPECT image at V2 (**Figure 3**).

Subject 6-8 were patients with mild COPD. They can be treated as C1 of current smokers at all visits given that they had marginal emphysema and fSAD as well as preserved lung function (13), although Subject 7 was located at the boundary of C1 and C3 at V0 and V1. They showed homogeneous TC distributions and no clear hot spot on the SPECT images for all visits (**Figure 3**).

Our findings indicate that fSAD was the primary factor determining both disease stage and the heterogeneity of TC distribution, and it was the most significant imaging variable distinguishing the clusters. Furthermore, the clusters appeared to provide a more sensitive representation of lung ventilation patterns compared to GOLD stages.

### Association between SPECT and qCT Variables

CV_Total_ and TC_Max_ demonstrated strong correlations with fSAD%_Total_ (CV_Total_: r=0.90; TC_Max_: r=0.86) and Emph%_Total_ (CV_Total_: r=0.71; TC_Max_: r=0.77) on cross-sectional data, meaning that fSAD and emphysema were associated with heterogeneity of lung ventilation. Compared to emphysema, fSAD was found to have a stronger association with the heterogeneity of lung ventilation. To further interpret and complement the aforementioned findings, a cross-lagged panel analysis study (see **Supplementary Material**) was conducted to examine the causal relationship between fSAD and heterogeneity of lung ventilation. The analysis indicated that fSAD is the cause of heterogeneity of lung ventilation. In COPD, narrowing of small airways leads to higher airflow resistance and greater particle deposition in the lungs (32–34). As a result, subjects with greater fSAD%_Total_ were observed to have localized areas of increased TC activity, or ‘hot spots’. Regarding the longitudinal data collected over two years, it was observed that there were strong correlations between TC_Max_ and D_h,sLUL_* (r=-0.74) as well as 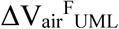 (r=0.75) (**Figure 5**). These findings suggest that reduced diameters of segmental airways and imbalanced ventilation between the upper and lower lobes may also play a role in the development of TC hot spots. Upon visualizing the segmental airways in the LUL of Subject 3 and 5, there existed constricted branches at V2 and V0, respectively (**Figure 7**). This finding reveals the negative correlations between the changes of D_h,sLUL_* and the changes of SPECT ventilation images. This observation is in line with a previous study that identified D_h,sLUL_* as a significant discriminant qCT variable for clusters of former smokers (14). Since changes of TC_Max_ over two years were negatively correlated with those of fSAD% (r=-0.70) and Emph% (r=-0.75), changes in D_h,sLUL_* and 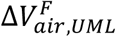 could moderate the correlation between changes in TC_Max_ and changes in fSAD%_Total_ and Emph%_Total_ when the disease stages remained relatively stable over time. This suggests that the formation of hot spots in individuals with COPD could be influenced by alterations in the segmental airways located in the left upper lobe, as well as changes in the distribution of airflow between the upper and lower lobes, in addition to factors such as fSAD and emphysema. Because of the small sample size and the minimal changes observed in SPECT and qCT variables between visits, further investigation with a larger sample size is necessary to validate the correlations found in the longitudinal data.

**Figure 7.**
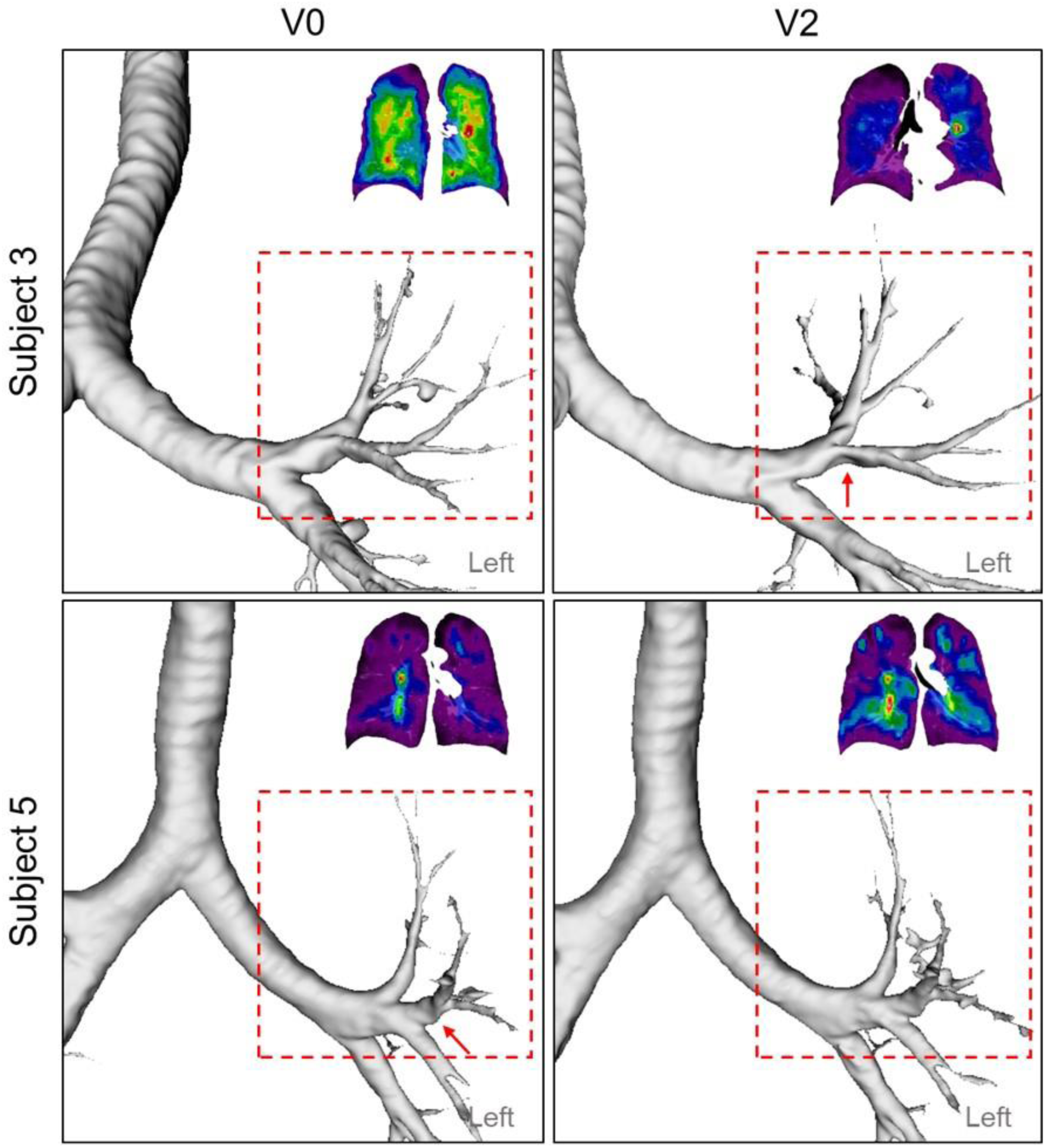
A constricted segmental airway in LUL (red arrow) of Subject 3 at V2, which may contribute to the TC hot spot found in LUL. In addition, a constricted segmental airway in LUL (red arrow) of Subject 5 at V0 resolved at V2, which may increase the homogeneity of ventilation.

This study was subject to several limitations. Firstly, the spatial resolution of SPECT images, which was 3.985 mm, may not be as precise as that of CT scans, which had a resolution of 0.5-0.7 mm. Consequently, there may be some inaccuracies in the quantification of SPECT images in certain regions (such as instances where the tracer may appear outside of the lung on CT images). Furthermore, the radiation exposure associated with SPECT is not insignificant, and its widespread availability is limited. An alternative avenue for this study involves lung ventilation derived from the XeMRI technique. This approach has been demonstrated to exhibit a high correlation with ventilation SPECT (18). Secondly, the sample size was relatively small, and thus increasing the size of the sample may be necessary to obtain more robust and reliable results. Furthermore, the fluctuations of J_Total_ between visits observed in Subject 1 and 7 may cause the unstable cluster memberships when the subjects were located near the boundaries of two clusters, even though there was no clear disease progression or recovery. These fluctuations of J_Total_ might be related to the variations of lung volume control during the CT scans. Lastly, it should be noted that the subjects in this study effectively managed their disease for a period of two years in terms of severity, and no significant changes in lung function or imaging were observed during this time. Thus, in order to obtain a clearer understanding of the progression of COPD, it may be necessary to extend the interval between visits beyond the two-year period evaluated in this study.

## CONCLUSION

To summarize, our analysis revealed three distinct categories of subjects: those with severe COPD (Subjects 2 and 5), those with moderate COPD (Subjects 1, 3, and 4), and those who exhibited mild symptoms or were at-risk for developing COPD (Subjects 6, 7, and 8). Interestingly, all subjects within each category displayed similar patterns of ventilation as observed in the SPECT images. Our findings suggest that fSAD could be a crucial factor that contributes to the heterogeneous ventilation observed in patients with COPD. Furthermore, the formation of hot spots in patients with COPD could potentially be influenced by alterations in the segmental airways located in the left upper lobe, along with modifications in the distribution of airflow between the upper and lower lobes, as well as the degree of fSAD and emphysema. Further investigation of fSAD could lead to a better understanding of the diverse characteristics of COPD, improved identification of COPD phenotypes, and enhanced knowledge of the progression of the disease.

## Data Availability

The data generated or analyzed during this study are included in this published article and its supplementary information files. The imaging datasets analyzed during the current study are not publicly available due privacy concern but are available from the corresponding author on reasonable request.

## DECLARATIONS

### Ethics Approval and Consent to Participate

This study (Title: Integrative lung model Y3-Y5) complies with the Declaration of Helsinki. It was approved by the institutional review board of the University of Iowa and informed consent was obtained before the study to have individual patient data published. The manuscript and the results were presented in a way that the patients cannot be identified.

### Consent for Publication

All authors approve the version to be published.

### Competing Interests

There is no conflict of interest for all authors.

### Funding/Support

Supports for this study were provided, in part, by NIH grants R01-HL168116, U01-HL114494, R01-HL112986, S10-RR022421, T32-HL-144461, and ED grant P116S210005.

### Author’s Contributions

All authors made a significant contribution to the work reported, whether that is in the conception, study design, execution, acquisition of data, analysis and interpretation, or in all these areas; took part in drafting, revising or critically reviewing the article; gave final approval of the version to be published; have agreed on the journal to which the article has been submitted; and agree to be accountable for all aspects of the work.

## Notes

### Competing Interest Statement

The authors have declared no competing interest.

### Author Declarations

This study (Title: Integrative lung model Y3-Y5) was approved by the institutional review board of the University of Iowa and informed consent was obtained before the study to have individual patient data published.

