## Supplementary Material for "Exploratory Study on COPD Phenotypes and their Progression: Integrating SPECT and qCT Imaging Analysis"

**Table S1.** Demographic data and PFT results of all the subjects at three visits. $R_{aw}$ is the airway resistance measured by plethysmography.

| | Gender | Age (yrs.) | BMI | Wight (kg) | Height (cm) | FEV1% predicted | FEV1/FVC | GOLD Stage | TLC (L) | FRC (L) | RV (L) | $R_{aw}$ (cmH2O/L/Sec) |
|---|---|---|---|---|---|---|---|---|---|---|---|---|
| **V0** | | | | | | | | | | | | |
| Subj. 1 | male | 71-80 | 31.6 | 108.0 | 185 | 73 | 53 | 2 | 7.48 | 5.03 | 2.98 | 1.18 |
| Subj. 2 | male | 81-90 | 22.9 | 65.5 | 169 | 50 | 28 | 2 | 7.91 | 6.11 | 3.88 | 1.31 |
| Subj. 3 | male | 51-60 | 25.6 | 88.6 | 186 | 63 | 51 | 2 | 7.21 | 3.83 | 2.24 | 1.04 |
| Subj. 4 | male | 51-60 | 21.4 | 61.1 | 169 | 73 | 56 | 2 | 6.93 | 4.73 | 2.81 | 1.27 |
| Subj. 5 | male | 51-60 | 31.6 | 85.0 | 164 | 36 | 28 | 3 | 6.79 | 4.79 | 3.24 | 1.75 |
| Subj. 6 | male | 51-60 | 29.3 | 92.8 | 178 | 83 | 59 | 1 | 6.76 | 3.71 | 1.22 | 1.26 |
| Subj. 7 | female | 51-60 | 27.0 | 75.4 | 167 | 77 | 71 | 0 | 5.07 | 2.63 | 1.99 | 1.34 |
| Subj. 8 | female | 61-70 | 35.5 | 91.0 | 160 | 88 | 77 | 0 | 4.23 | 1.67 | 1.53 | 1.76 |
| **Mean±SD** | | | 28.1 ± 4.5 | 83.4 ± 14.4 | 172.3 ± 9.0 | 67.9 ± 16.3 | 52.9 ± 16.6 | | 6.5 ± 1.2 | 4.1 ± 1.3 | 2.5 ± 0.8 | 1.4 ± 0.2 |
| **V1** | | | | | | | | | | | | |
| | **Gender** | **Age (yrs.)** | **BMI** | **Wight (kg)** | **Height (cm)** | **FEV1% predicted** | **FEV1/FVC** | **GOLD Stage** | **TLC (L)** | **FRC (L)** | **RV (L)** | **$R_{aw}$ (cmH2O/L/Sec)** |
| Subj. 1 | male | | 28.7 | 98.2 | 185 | 74 | 56 | 2 | 7.76 | 5.71 | 3.54 | 1.12 |
| Subj. 2 | male | | 26.0 | 74.3 | 169 | 39 | 26 | 3 | 7.76 | 6.02 | 4.4 | 1.4 |
| Subj. 3 | male | | 26.2 | 90.5 | 186 | 61 | 51 | 2 | 7.62 | 3.7 | 2.75 | 1.04 |
| Subj. 4 | male | | 20.5 | 58.6 | 169 | 68 | 58 | 2 | 7.2 | 4.71 | 3.12 | 1.25 |
| Subj. 5 | male | | 30.6 | 82.4 | 164 | 31 | 25 | 3 | 7.1 | 4.94 | 3.73 | 1.71 |
| Subj. 6 | male | | 29.3 | 92.8 | 178 | 78 | 58 | 2 | 7.37 | 3.71 | 2.06 | 1.26 |
| Subj. 7 | female | | 26.8 | 75.7 | 168 | 75 | 71 | 0 | 4.69 | 2.84 | 1.68 | 1.34 |
| Subj. 8 | female | | 33.6 | 86 | 160 | 97 | 78 | 0 | 4.23 | 2.21 | 1.34 | 1.69 |
| **Mean±SD** | | | 27.7 ± 3.6 | 172.4 ± 9.0 | 82.3 ± 11.8 | 65.4 ± 20.1 | 52.9 ± 17.8 | | 6.7 ± 1.3 | 4.2 ± 1.3 | 2.8 ± 1.0 | 1.4 ± 0.2 |
| **V2** | | | | | | | | | | | | |
| | **Gender** | **Age (yrs.)** | **BMI** | **Wight (kg)** | **Height (cm)** | **FEV1% predicted** | **FEV1/FVC** | **GOLD Stage** | **TLC (L)** | **FRC (L)** | **RV (L)** | **$R_{aw}$ (cmH2O/L/Sec)** |
| Subj. 1 | male | | 28.7 | 98.2 | 185 | 76 | 53 | 2 | 7.6 | 5.29 | 3.09 | 1.12 |
| Subj. 2 | male | | 26.0 | 74.3 | 169 | 51 | 29 | 2 | 8.11 | 6.31 | 4.66 | 1.4 |
| Subj. 3 | male | | 25.4 | 87.8 | 186 | 62 | 51 | 2 | 7.6 | 3.82 | 2.68 | 1.06 |
| Subj. 4 | male | | 20.8 | 59.3 | 169 | 78 | 60 | 2 | 7.05 | 4.61 | 3.26 | 1.24 |
| Subj. 5 | male | | 30.6 | 82.4 | 164 | 33 | 28 | 3 | 7.17 | 5.21 | 3.76 | 1.71 |
| Subj. 6 | male | | 29.3 | 92.8 | 178 | 81 | 59 | 1 | 6.89 | 3.07 | 1.63 | NA |

| | | | | | | | | | | | | |
|---|---|---|---|---|---|---|---|---|---|---|---|---|
| Subj. 7 | female | | 27.0 | 75.4 | 167 | 73 | 71 | 0 | 4.98 | 2.7 | 2.1 | 1.31 |
| Subj. 8 | female | | 33.6 | 86 | 160 | 97 | 78 | 0 | 4.33 | 1.93 | 1.49 | NA |
| **Mean±SD** | | | 27.7 ± 3.6 | 172.3 ± 9.0 | 82.0 ± 11.4 | 68.9 ± 18.5 | 53.6 ± 16.7 | | 6.7 ± 1.3 | 4.1 ± 1.4 | 2.8 ± 1.0 | 1.3 ± 0.2 |

**Table S2.** TC% and $\Delta V_{air}^{F}$ of each lobe at each visit.

| ID | TC% | Lobe | Visit | $\Delta V_{air}^{F}$ |
|---|---|---|---|---|
| Subj.3 | 0.192 | LLL | V0 | 0.154 |
| Subj.4 | 0.281 | LLL | V0 | 0.296 |
| Subj.5 | 0.183 | LLL | V0 | 0.149 |
| Subj.6 | 0.229 | LLL | V0 | 0.153 |
| Subj.7 | 0.284 | LLL | V0 | 0.215 |
| Subj.8 | 0.295 | LLL | V0 | 0.227 |
| Subj.3 | 0.272 | LUL | V0 | 0.291 |
| Subj.4 | 0.215 | LUL | V0 | 0.200 |
| Subj.5 | 0.316 | LUL | V0 | 0.336 |
| Subj.6 | 0.258 | LUL | V0 | 0.273 |
| Subj.7 | 0.183 | LUL | V0 | 0.217 |
| Subj.8 | 0.186 | LUL | V0 | 0.221 |
| Subj.3 | 0.180 | RLL | V0 | 0.165 |
| Subj.4 | 0.283 | RLL | V0 | 0.268 |
| Subj.5 | 0.191 | RLL | V0 | 0.172 |
| Subj.6 | 0.237 | RLL | V0 | 0.035 |
| Subj.7 | 0.291 | RLL | V0 | 0.223 |
| Subj.8 | 0.290 | RLL | V0 | 0.237 |
| Subj.3 | 0.037 | RML | V0 | 0.035 |
| Subj.4 | 0.059 | RML | V0 | 0.066 |
| Subj.5 | 0.075 | RML | V0 | 0.107 |
| Subj.6 | 0.055 | RML | V0 | 0.427 |
| Subj.7 | 0.049 | RML | V0 | 0.064 |
| Subj.8 | 0.062 | RML | V0 | 0.084 |
| Subj.3 | 0.319 | RUL | V0 | 0.355 |
| Subj.4 | 0.163 | RUL | V0 | 0.170 |
| Subj.5 | 0.234 | RUL | V0 | 0.236 |
| Subj.6 | 0.222 | RUL | V0 | 0.112 |
| Subj.7 | 0.193 | RUL | V0 | 0.281 |
| Subj.8 | 0.167 | RUL | V0 | 0.230 |
| Subj.1 | 0.242 | LUL | V1 | 0.178 |
| Subj.2 | 0.287 | LUL | V1 | 0.305 |
| Subj.3 | 0.311 | LUL | V1 | 0.283 |
| Subj.4 | 0.193 | LUL | V1 | 0.212 |
| Subj.5 | 0.330 | LUL | V1 | 0.311 |
| Subj.6 | 0.259 | LUL | V1 | 0.258 |
| Subj.7 | 0.183 | LUL | V1 | 0.186 |
| Subj.8 | 0.250 | LUL | V1 | 0.180 |
| Subj.1 | 0.151 | LLL | V1 | 0.281 |
| Subj.2 | 0.170 | LLL | V1 | 0.119 |
| Subj.3 | 0.126 | LLL | V1 | 0.178 |
| Subj.4 | 0.345 | LLL | V1 | 0.268 |
| Subj.5 | 0.144 | LLL | V1 | 0.176 |

| | | | | |
|---|---|---|---|---|
| Subj.6 | 0.185 | LLL | V1 | 0.223 |
| Subj.7 | 0.244 | LLL | V1 | 0.287 |
| Subj.8 | 0.182 | LLL | V1 | 0.297 |
| Subj.1 | 0.203 | RUL | V1 | 0.139 |
| Subj.2 | 0.306 | RUL | V1 | 0.322 |
| Subj.3 | 0.417 | RUL | V1 | 0.331 |
| Subj.4 | 0.169 | RUL | V1 | 0.173 |
| Subj.5 | 0.211 | RUL | V1 | 0.254 |
| Subj.6 | 0.258 | RUL | V1 | 0.200 |
| Subj.7 | 0.237 | RUL | V1 | 0.187 |
| Subj.8 | 0.258 | RUL | V1 | 0.170 |
| Subj.1 | 0.138 | RML | V1 | 0.109 |
| Subj.2 | 0.110 | RML | V1 | 0.121 |
| Subj.3 | 0.026 | RML | V1 | 0.038 |
| Subj.4 | 0.053 | RML | V1 | 0.064 |
| Subj.5 | 0.103 | RML | V1 | 0.073 |
| Subj.6 | 0.084 | RML | V1 | 0.095 |
| Subj.7 | 0.054 | RML | V1 | 0.049 |
| Subj.8 | 0.086 | RML | V1 | 0.063 |
| Subj.1 | 0.266 | RLL | V1 | 0.294 |
| Subj.2 | 0.128 | RLL | V1 | 0.133 |
| Subj.3 | 0.121 | RLL | V1 | 0.171 |
| Subj.4 | 0.240 | RLL | V1 | 0.285 |
| Subj.5 | 0.212 | RLL | V1 | 0.186 |
| Subj.6 | 0.215 | RLL | V1 | 0.224 |
| Subj.7 | 0.282 | RLL | V1 | 0.292 |
| Subj.8 | 0.225 | RLL | V1 | 0.291 |
| Subj.1 | 0.271 | LUL | V2 | 0.192 |
| Subj.2 | 0.273 | LUL | V2 | 0.337 |
| Subj.3 | 0.365 | LUL | V2 | 0.312 |
| Subj.4 | 0.209 | LUL | V2 | 0.235 |
| Subj.5 | 0.343 | LUL | V2 | 0.300 |
| Subj.6 | 0.286 | LUL | V2 | 0.259 |
| Subj.7 | 0.205 | LUL | V2 | 0.202 |
| Subj.8 | 0.246 | LUL | V2 | 0.195 |
| Subj.1 | 0.127 | LLL | V2 | 0.267 |
| Subj.2 | 0.193 | LLL | V2 | 0.095 |
| Subj.3 | 0.113 | LLL | V2 | 0.109 |
| Subj.4 | 0.372 | LLL | V2 | 0.245 |
| Subj.5 | 0.153 | LLL | V2 | 0.196 |
| Subj.6 | 0.130 | LLL | V2 | 0.221 |
| Subj.7 | 0.231 | LLL | V2 | 0.260 |
| Subj.8 | 0.194 | LLL | V2 | 0.283 |
| Subj.1 | 0.224 | RUL | V2 | 0.157 |
| Subj.2 | 0.279 | RUL | V2 | 0.319 |
| Subj.3 | 0.401 | RUL | V2 | 0.411 |

| | | | | |
|---|---|---|---|---|
| Subj.4 | 0.151 | RUL | V2 | 0.179 |
| Subj.5 | 0.220 | RUL | V2 | 0.246 |
| Subj.6 | 0.293 | RUL | V2 | 0.241 |
| Subj.7 | 0.267 | RUL | V2 | 0.210 |
| Subj.8 | 0.235 | RUL | V2 | 0.163 |
| Subj.1 | 0.161 | RML | V2 | 0.112 |
| Subj.2 | 0.113 | RML | V2 | 0.124 |
| Subj.3 | 0.020 | RML | V2 | 0.034 |
| Subj.4 | 0.047 | RML | V2 | 0.063 |
| Subj.5 | 0.090 | RML | V2 | 0.066 |
| Subj.6 | 0.121 | RML | V2 | 0.069 |
| Subj.7 | 0.058 | RML | V2 | 0.057 |
| Subj.8 | 0.093 | RML | V2 | 0.055 |
| Subj.1 | 0.217 | RLL | V2 | 0.271 |
| Subj.2 | 0.141 | RLL | V2 | 0.124 |
| Subj.3 | 0.100 | RLL | V2 | 0.134 |
| Subj.4 | 0.220 | RLL | V2 | 0.279 |
| Subj.5 | 0.194 | RLL | V2 | 0.192 |
| Subj.6 | 0.169 | RLL | V2 | 0.211 |
| Subj.7 | 0.238 | RLL | V2 | 0.271 |
| Subj.8 | 0.233 | RLL | V2 | 0.304 |

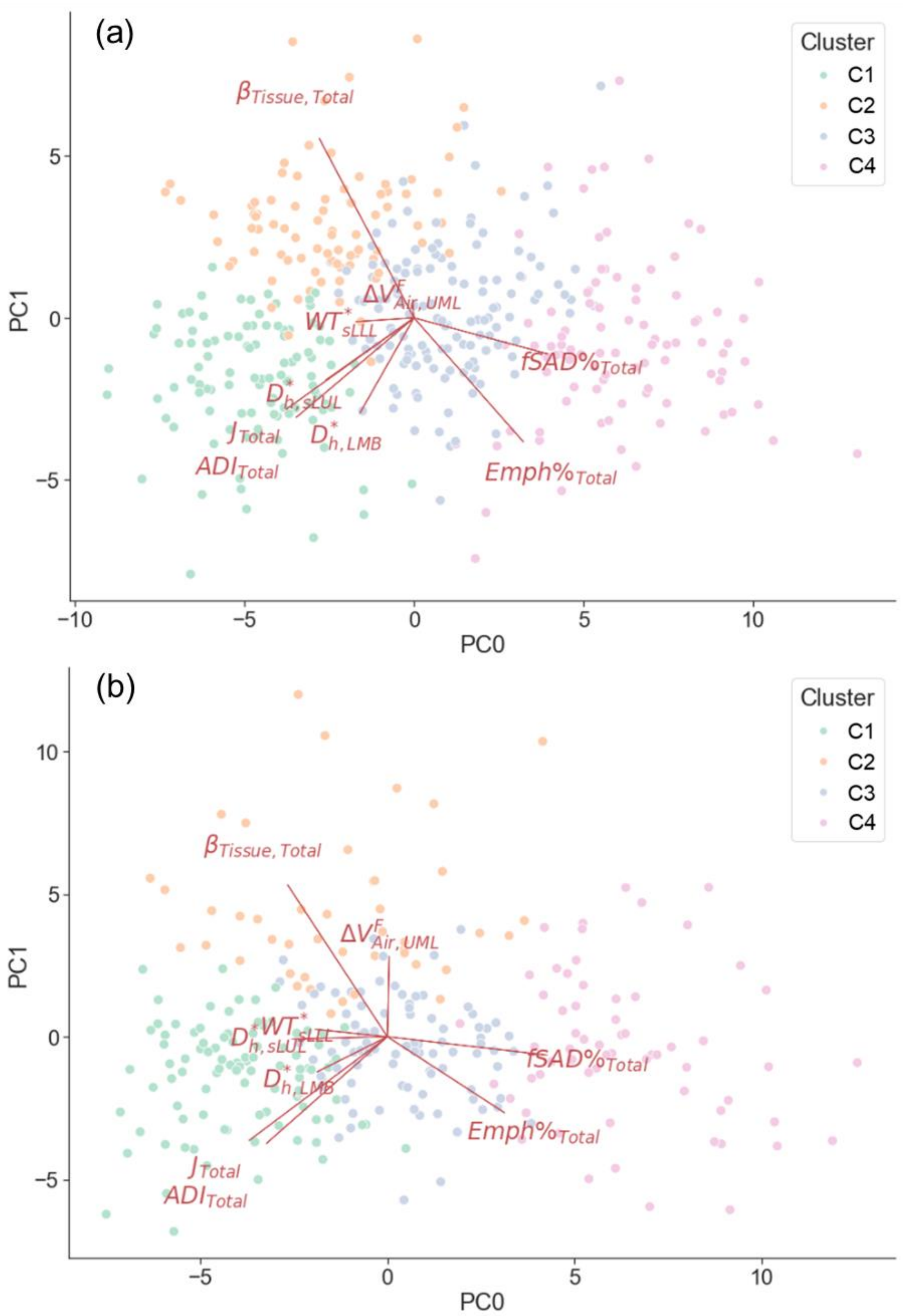


**Figure S1.** Biplots of the (a) former and (b) current smoker's data. The scattered points were the PC scores of observations and the arrows demonstrated the contributions and directions of the selected imaging metrics to the PC0 and PC1.

**Supplementary Content 1: Multiscale qCT Biomarkers**

The multiscale qCT biomarkers included lung structural and functional variables. The structural variables, describing the regional alterations of lung structures, included bifurcation angles (θ) between the children branches of trachea and right main bronchus (RMB), airway circularity (Cr), normalized airway wall thickness ($WT^*$), and normalized airway hydraulic diameter ($D_h^*$). Decreased θ and decreased Cr were found to be associated with airflow limitation (1) and increased functional small airway disease (2), respectively. The dimensions of wall thickness and hydraulic diameter were normalized by predicted trachea wall thickness and hydraulic diameter from healthy controls to eliminate inter-subject variability due to sex, age, and height (3). $WT^*$ and $D_h^*$ reveal the effects of wall thickening and luminal narrowing on airway obstructions caused by inflammation and hyper-responsiveness, respectively. The functional variables, capturing the regional alterations of lung functions, included fractional air volume change ($\Delta V_{air}^{F}$), determinant of Jacobian matrix (J), anisotropic deformation index (ADI), fraction-based small airways disease (fSAD%), fraction-based emphysema (Emph%), and tissue fraction at TLC ($\beta_{tissue}$). $\Delta V_{air}^{F}$ was quantified by the ratio of the lobar air-volume change to the whole lung air-volume change. The ratio of the air-volume change of the upper lobes to the air-volume change of the middle and lower lobes combined ($\Delta V_{air,UML}^{F}$) was also calculated. Jacobian (J) is a measure of local specific volume assessing the functional capacity of lung tissue. ADI is a measure of the magnitude of anisotropic deformation (4). Emph% and fSAD% are used to quantify the emphysematous lung tissue destruction and the extent of small airway narrowing or closure, respectively (5). $\beta_{tissue}$ indicates the proportion of tissue volume for detection of tissue destruction.

**Supplementary Content 2: Abbreviation List of qCT Variables**

The region where the qCT variable was measured was specified as the subscript of the variable, namely $\{\text{Variable}\}_{\{\text{Region}\}}$.

***Regions:***

LUL: Left upper lobe

LLL: Left lower lobe

RUL: Right upper lobe

RML: Right middle lobe

RLL: Right lower lobe

Total: Total lung

RMB: Right main bronchus

LMB: Left main bronchus

BronInt: Right intermediate bronchus

TriLLB: Trifurcation of left lower lobe

sLUL: Sub-grouped segmental airways of left upper lobe

sLLL: Sub-grouped segmental airways of left lower lobe

sRUL: Sub-grouped segmental airways of right upper lobe

sRML: Sub-grouped segmental airways of right middle lobe

sRLL: Sub-grouped segmental airways of right lower lobe

***Structural Variables:***

$\theta$: Bifurcation angles between the children branches of trachea and RMB

Cr: Airway circularity

$WT^*$: Normalized airway wall thickness

$D_h^*$: Normalized airway hydraulic diameter

***Functional Variables:***

$\Delta V_{air}^{F}$: Fractional air volume change

J: Determinant of Jacobian matrix

ADI: Anisotropic deformation index

fSAD%: Fraction-based small airways disease

Emph%: Fraction-based emphysema

$\beta_{tissue}$: Tissue fraction at TLC

$\Delta V_{air,UML}^{F}$: The ratio of the air-volume change of the upper lobes to the air-volume change of the middle and lower lobes combined

**Supplementary Content 3: A Post-hoc Study**

A post-hoc study was conducted to establish the causal relationship between fSAD and heterogeneity of lung ventilation. Cross-lagged panel analysis, which is commonly used to infer the direction and strength of a relationship between two variables repeatedly measured at different time points, was employed (6). The two imaging variables of $fSAD\%_{Total}$ and $CV_{Total}$ at V1 and V2 were chosen for analysis since they were strongly correlated at all visits. The type one error rate (α) was set at 0.05. As demonstrated in **Figure S2**, two synchronous correlations ($r_{fSAD1,CV1}$ and $r_{fSAD2,CV2}$) and two stability correlations ($r_{CV1,CV2}$ and $r_{fSAD1,fSAD2}$) were both significantly greater than zero, indicating that the assumptions of synchronicity and stationarity were not violated. Regarding the cross-lagged correlations ($r_{fSAD1,CV2}$ and $r_{CV1,fSAD2}$), $r_{fSAD1,CV2}$ was significantly greater than zero while $r_{CV1,fSAD2}$ was not, suggesting that fSAD is the cause of heterogeneity of lung ventilation. Note that the cross-lagged correlations were partial correlations which the contributions of the stability correlations had been partialled out.

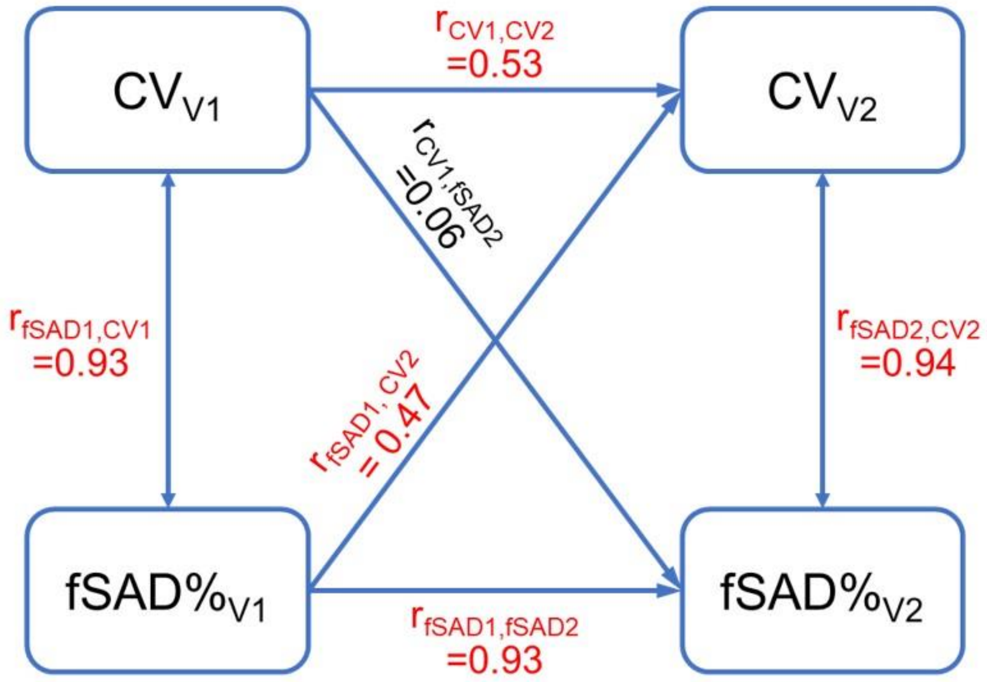


**Figure S2.** The cross-lagged panel analysis estimated a total of six correlations. The correlations colored in red were significantly greater than zero. The result that $r_{fSAD1,CV2}$ was greater than zero suggested that fSAD cause the heterogeneity of lung ventilation.
